# Contextual and Landscape Analysis of Team-Based Care Practices for Hypertension Management in Ghana: A Cross-Sectional Nationwide Study

**DOI:** 10.1101/2025.08.01.25332564

**Authors:** Thomas Hinneh, Dzifa Ahadzi, Samuel Byirigiro, Faith E. Metlock, Oluwabunmi Ogungbe, Cheryl R. Dennison Himmelfarb, Lawrence Appel, Fred Stephen Sarfo, Yvonne Commodore-Mensah

**Affiliations:** Johns Hopkins School of Nursing, Baltimore, Maryland, United States; Tamale Teaching Hospital, Tamale, Ghana; Vanderbilt University School of Nursing, NA, Tennessee; Johns Hopkins ProHealth Clinical Research, Baltimore, Maryland, United States; Komfo Anokye Teaching Hospital, Kumasi, Ghana

**Keywords:** Team based care, Hypertension, Task shifting, healthcare systems, non-physician healthcare workers LMICs, Ghana

## Abstract

**Introduction:** The World Health Organization recommends team-based care (TBC) for hypertension control, particularly in low-resourced settings. This study assessed current practices, task distribution, and perspectives on a team-based approach to hypertension management in Ghana.

**Methods:** In this cross-sectional study, we used convenience sampling to disseminate an online Resolve to Save Lives Survey (RTSL) to healthcare workers (HCWs) involved in hypertension management. Hypertension task-complexity was conceptualized (administrative, basic, and advanced clinical tasks) based on the Team-Based Hypertension Care conceptual framework and stratified by HCWs and facility-level characteristics.

**Results:** Among 345 HCWs, the mean age was 34 (±6.3), 58% were males, and 48% practiced in urban centers. Clinical task performance varies by setting, level of care, and education. Most administrative tasks were performed by non-clinicians (69%) and community health workers (14%). Basic clinical tasks were team-based, shared among nurses, pharmacists, and physician assistants. Nearly all advanced tasks were handled by physicians (28%) and physician assistants (43%). Major barriers to TBC included limited scope-of-practice laws (79%), inadequate training of non-physician workers (74%), opposition by physicians (62%), resistance by patients (57%), and opposition by nurses (43%). Utilization of treatment algorithms (96%), m-health technology (93%), and adequate HCW compensation (79%) were notable facilitators.

**Conclusions:** With dire shortages of physicians, strengthening the capacity of non- physician HCWs to perform advanced clinical tasks is essential for effective hypertension care in Ghana. Policies are needed to support training, expanded scope of practice, and regulatory reform to advance team-based care.

## INTRODUCTION

Hypertension is a major global public health challenge and the leading modifiable risk factor for cardiovascular disease and premature death[1]. More than 1.3 billion people globally are affected by hypertension, with the majority living in low- and middle-income countries (LMICs)[1,2]. Despite the availability of effective, low-cost treatments, blood pressure control remains suboptimal, with less than 30% of people achieving control worldwide, with the worst estimates in LMICs [2]. In Ghana, for example, the physician-to-patient ratio is approximately 1 to 6,400, while the nurse-to-patient ratio is about 1 to 700. The ratio is even worse in deprived regions [3,4]. This gap presents an important opportunity to involve nurses and other clinicians more actively in hypertension care, which can improve patient engagement and the overall quality of care. To support effective hypertension management, the World Health Organization (WHO) recommends team-based care (TBC) as a core strategy, especially in settings with limited resources [5].

Team-based care refers to collaborative, coordinated patient management involving two or more HCWs, such as physicians, nurses, pharmacists, and other HCWs[6]. Systematic reviews and meta-analyses have shown evidence that care models enabling non-physician HCWs to take on advanced clinical tasks, such as titrating antihypertensive medications, are effective and cost-efficient [7–10]. More importantly, the applicability of evidence extends beyond developed countries to LMICs [11–13].

In Africa, the burden of hypertension continues to rise. Yet, health systems are often ill- equipped with significant gaps across the entire continuum of hypertension care, with resulting debilitating impacts including hypertension-related complications and avoidable deaths[14,15]. Although efforts are underway to adapt TBC strategies to local contexts through task-sharing, implementation remains inconsistent and often ineffective due to policy restrictions and a lack of sustainable funding to drive commitment [16].

The hypertension care cascade in Ghana reflects critical health system gaps: fewer than 35% of adults with hypertension are aware of their condition, and only about one in four achieve blood pressure control [2]. Earlier clinical-level data have suggested that more than 65% of patients attending hypertension clinics across health facilities in Ghana had uncontrolled blood pressure, suggesting a decline in HTN control rates [17]. Although several components of the WHO-HEARTS package, such as healthy-lifestyle counseling, the availability of evidence-based treatment protocols, access to essential medicines and technology, and systems for monitoring, have been incorporated into mainstream hypertension care, the components related to team-based care and risk-based cardiovascular disease assessment are poorly integrated into care in Ghana[5]. To strategically address these systemic gaps and lay the foundation for a guided implementation of team-based hypertension management, it is essential to assess the scope of team-based hypertension care to inform effective implementation efforts.

This study examines current TBC practices for hypertension management among HCWs and their perspectives on perceived barriers and facilitators to TBC implementation.

## METHODS

### Design

We designed a cross-sectional survey-based study consisting of an online survey targeting HCWs involved in hypertension care in Ghana. The survey has been designed to gather information on team-based care practices in hypertension management and has been used previously in other LMICs; its development and methodology are described[18]. However, to ensure contextual feasibility, the survey was piloted with five HCWs and two non- communicable disease research experts.

The study conduct and findings reporting are aligned with the Strengthening the Reporting of Observational Studies in Epidemiology (STROBE) guideline for cross-sectional studies.

### Data Collection

The survey was administered in all 16 administrative regions of Ghana between June 2024 and December 2024. The survey was distributed in English to various cadres of HCWs across all 16 administrative regions of Ghana. Eligible participants included nurses, physicians, pharmacists, and community health officers. The survey aimed to understand perspectives on team-based management of hypertension, including the barriers and facilitators. A convenience sampling approach was used with a secure, encrypted link on REDCap distributed through professional networks and healthcare facility contacts in Ghana. Data on sociodemographic facility-level characteristics and task distribution were also collected. In cases where the selected representative could not complete the survey, an alternative individual from the same organization or program was recommended. Participation was voluntary, and all respondents provided informed consent before survey completion (**Online Supplemental Material** Appendix 1).

### Study Population

The target respondents included frontline HCWs involved in hypertension care. No specific recruitment target was set, enabling us to mimic the natural distribution of the health workforce in Ghana.

### Ethics

This study was reviewed and approved by the Ghana Health Service Ethics Review Committee (Protocol ID: GHS-ERC026/03/23). All study procedures complied with the ethical standards of the institutional and national research committees and with the 1964 Helsinki Declaration and its later amendments. Participation in the study was voluntary. All participants were informed about the purpose of the study and provided electronic informed consent before initiating the survey.

### Statistical Analysis

Descriptive statistics were used to summarize participant characteristics. Continuous variables were expressed as means with standard deviations (SD), while categorical variables were presented as frequencies and percentages.

Hypertension management tasks were categorized using the Team-Based Hypertension Care conceptual framework, which the research team developed and previously applied in LMICs: (1) administrative tasks requiring minimal clinical training; (2) basic clinical tasks requiring moderate clinical judgment; and (3) advanced clinical tasks involving complex decision-making [18] (**Online Supplemental Figure 1).** Task distributions were stratified by HCW cadre, healthcare setting, and facility level, and visually presented using figures for easy interpretation. Pearson’s chi-square test or Fisher’s exact test was applied to examine differences in team-based care perceptions and task assignments across HCW groups, regions, and facility types. All statistical tests were two-sided, and a significant level of p < 0.05 was applied throughout the analysis. Survey responses were exported from REDCap and analyzed using Stata I/C version 16.1 (StataCorp, College Station, TX, USA).

## RESULTS

### Respondent Characteristics

A total of 345 HCWs involved in hypertension care in Ghana participated in the study. The mean age of the respondents was 34 years (SD ±6.3); 174 (58%) were men, and most were nursing professionals, 129 (45%) (**Table 1**). Nearly half, 142 (47%) of the respondents held some form of management role. Education levels varied among respondents: 134 (48%) held a bachelor’s degree, 66 (24%) had a post-secondary diploma, 53 (19%) held a master’s degree, 14 (5%) had a doctorate, and 10 (4%) reported vocational or other training. Most of the respondents worked in an urban setting, 134 (48%). The majority of the respondents, 87 (55%), worked at district/general hospitals, 42 (27%) at regional/tertiary hospitals, 26 (16%) at health centers, and 3 (2%) at polyclinics.

**Table 1:**
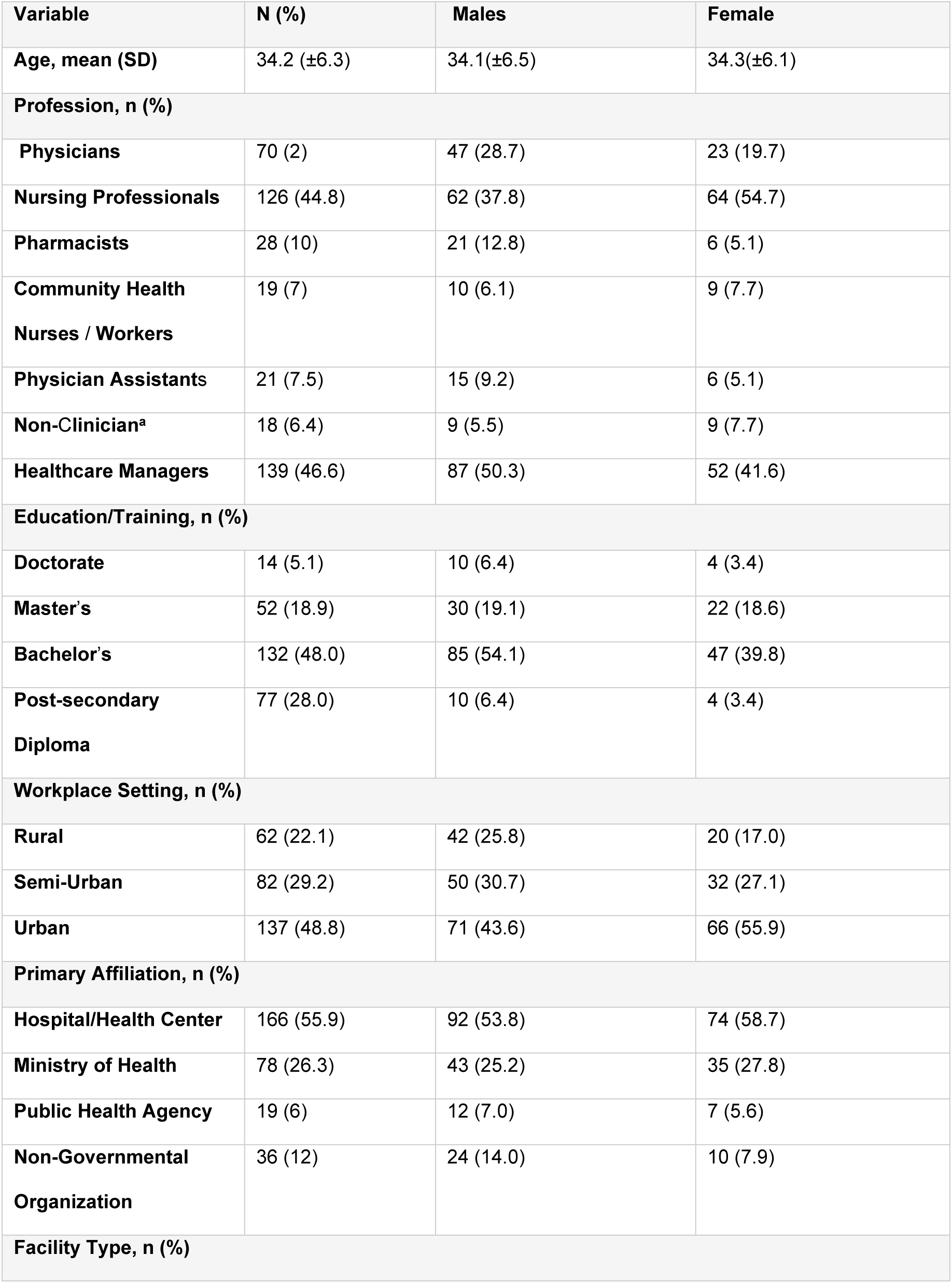
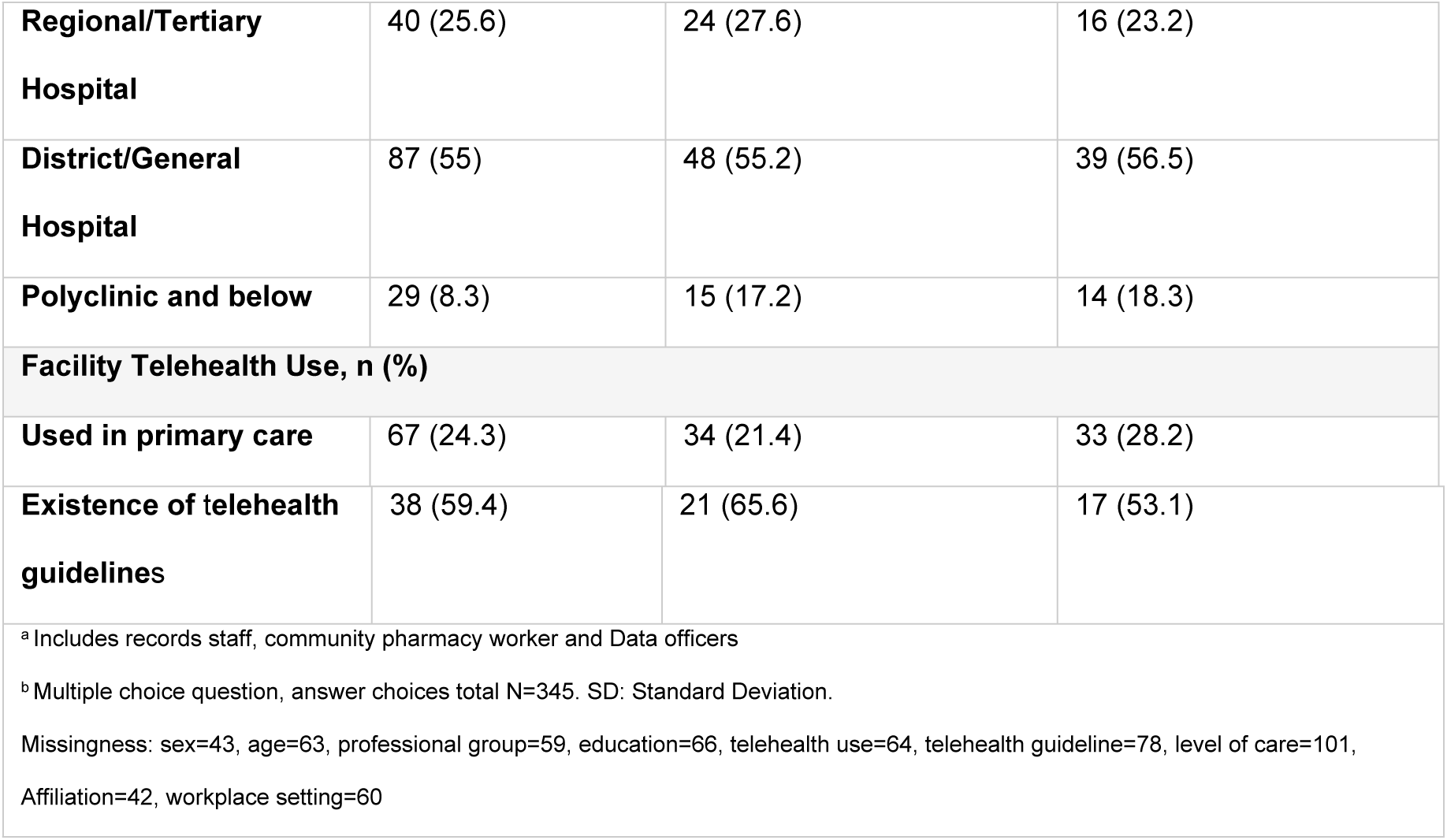
Characteristics of Respondents Involved in Hypertension Care in Ghana

| Variable | N (%) | Males | Female |
| --- | --- | --- | --- |
| Age, mean (SD) | 34.2 (±6.3) | 34.1(±6.5) | 34.3(±6.1) |
| **Profession, n (%)** |  |  |  |
| Physicians | 70 (2) | 47 (28.7) | 23 (19.7) |
| Nursing Professionals | 126 (44.8) | 62 (37.8) | 64 (54.7) |
| Pharmacists | 28 (10) | 21 (12.8) | 6 (5.1) |
| Community Health<br>Nurses / Workers | 19 (7) | 10 (6.1) | 9 (7.7) |
| Physician Assistants | 21 (7.5) | 15 (9.2) | 6 (5.1) |
| Non-Clinician<sup>a</sup> | 18 (6.4) | 9 (5.5) | 9 (7.7) |
| Healthcare Managers | 139 (46.6) | 87 (50.3) | 52 (41.6) |
| **Education/Training, n (%)** |  |  |  |
| Doctorate | 14 (5.1) | 10 (6.4) | 4 (3.4) |
| Master's | 52 (18.9) | 30 (19.1) | 22 (18.6) |
| Bachelor's | 132 (48.0) | 85 (54.1) | 47 (39.8) |
| Post-secondary<br>Diploma | 77 (28.0) | 10 (6.4) | 4 (3.4) |
| **Workplace Setting, n (%)** |  |  |  |
| Rural | 62 (22.1) | 42 (25.8) | 20 (17.0) |
| Semi-Urban | 82 (29.2) | 50 (30.7) | 32 (27.1) |
| Urban | 137 (48.8) | 71 (43.6) | 66 (55.9) |
| **Primary Affiliation, n (%)** |  |  |  |
| Hospital/Health Center | 166 (55.9) | 92 (53.8) | 74 (58.7) |
| Ministry of Health | 78 (26.3) | 43 (25.2) | 35 (27.8) |
| Public Health Agency | 19 (6) | 12 (7.0) | 7 (5.6) |
| Non-Governmental<br>Organization | 36 (12) | 24 (14.0) | 10 (7.9) |
| **Facility Type, n (%)** |  |  |  |
| **Regional/Tertiary Hospital** | 40 (25.6) | 24 (27.6) | 16 (23.2) |
| **District/General Hospital** | 87 (55) | 48 (55.2) | 39 (56.5) |
| **Polyclinic and below** | 29 (8.3) | 15 (17.2) | 14 (18.3) |
| **Facility Telehealth Use, n (%)** |  |  |  |
| **Used in primary care** | 67 (24.3) | 34 (21.4) | 33 (28.2) |
| **Existence of telehealth guidelines** | 38 (59.4) | 21 (65.6) | 17 (53.1) |
| <sup>a</sup> Includes records staff, community pharmacy worker and Data officers<br><sup>b</sup> Multiple choice question, answer choices total N=345. SD: Standard Deviation.<br>Missingness: sex=43, age=63, professional group=59, education=66, telehealth use=64, telehealth guideline=78, level of care=101, Affiliation=42, workplace setting=60 |  |  |  |

Telehealth use at the facility level was generally low, reported by only 67 respondents (24%). Telehealth use was highest in the Greater Accra and Bono regions (13%) each and lowest in the Northeast and Ahafo regions (2%). More than half, 38 (59%), indicated the availability of a telehealth guideline. No distinct differences were observed when HCWs and facility-level characteristics were stratified by region (**Online Supplemental Table 1**).

### Distribution of Healthcare Workers

There was a relatively even distribution of responses across all 16 regions of Ghana, reflecting the general distribution of HCWs in the country (**Online Supplemental Figure 1**). For example, 23% of physicians were based in the Greater Accra Region, followed by 10% in the Ashanti Region. Fewer physicians were located in newly created administrative regions such as the Bono East and Western North regions. Similarly, nurses were more evenly spread across regions, with 12% based in the Greater Accra region, followed by the Northern Region.

### Hypertension Task Distribution

Task assignments varied significantly by HCW. HCWs were asked to indicate which cadre was responsible for performing a specific hypertension-management task. Based on the hypertension team-based care conceptual framework, tasks were grouped into administrative, basic clinical, and advanced clinical categories. For administrative tasks, most responsibilities are handled by non-clinicians (69%), CHN/CHWs (14%), except for scheduling, which is primarily handled by prescribers (mostly physicians (33%) and physician assistants (39%). All clinicians were involved in performing basic clinical tasks, including physician assistants (35%), nurses (19%), pharmacists (28%), and physicians (2%). However, history taking and medication-related tasks, including prescribing and refilling medications, were handled mainly by physicians and physician assistants. Largely, HCWs operated within the boundaries of their core competencies. For instance, patient retrieval and medication delivery were performed primarily by CHN/CHWs (44%) and pharmacists (44%), respectively. However, advanced clinical tasks were handled mainly by physicians (43%) and physician assistants (28%) (**Figure 1**).

**Figure 1.**
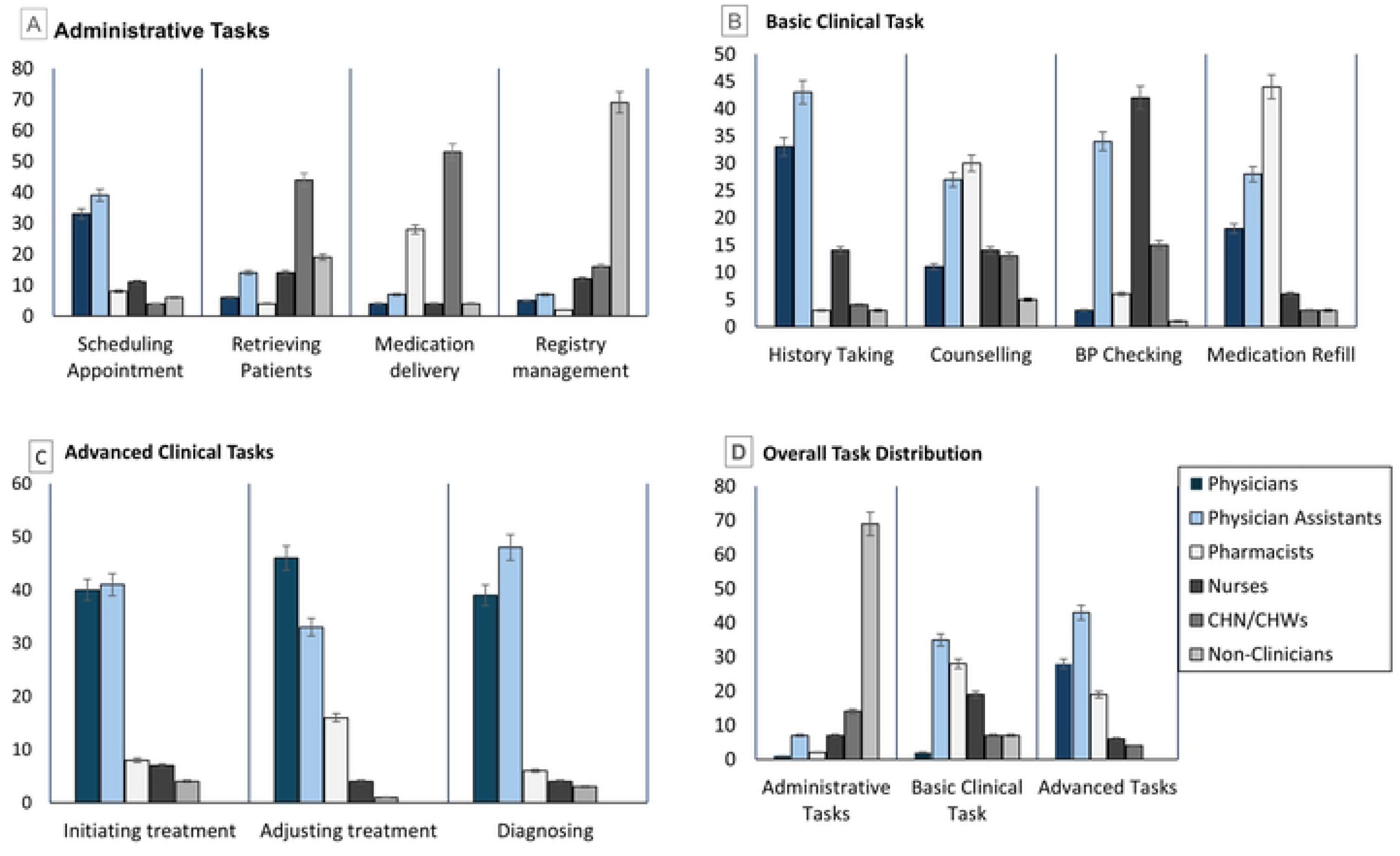
Hypertension task distribution based on the team-based hypertension care conceptual framework

When task distribution was stratified by health facility type and HCW demographics, administrative tasks were more commonly performed in district-level facilities and rural or semi-urban settings, with HCWs who held lower educational qualifications more likely to engage in these duties. Advanced clinical tasks were more frequently performed in urban healthcare settings (**Online Supplemental Figure 2**).

### Hypertension Treatment Schedule

Most of the HCWs (182, 62%) reported that a 30-day prescription was the standard duration for hypertension medication refills. Patients with controlled blood pressure (BP) were routinely seen monthly (126, 43.6%). In contrast, those with uncontrolled BP were often scheduled for follow-up visits more frequently, typically less than 30 days (240, 82.1%) (**Online Supplemental Figure 3)**

### Perspectives on Team-Based Hypertension Care

The perspectives of HCWs regarding team-based care were assessed in four thematic areas: patient outcomes and safety, workforce capacity and development, and health system strengthening. There was a consistently favorable perception among HCWs regarding the value of team-based care across these domains (**Figure 2**). A significant majority of HCWs, 93%, agreed that team-based care is necessary, with an even higher proportion, 98%, suggesting benefit in increasing the capacity for effective patient care, addressing physician shortage (78%), and improving healthcare coverage (96%). Regarding cost implications, 57% of HCWs felt that team-based care could reduce the cost of training HCWs and patient care costs (78%). About 74% of HCWs, however, indicated the need for financial compensation for additional responsibilities, and competency-based certification for HCWs involved in tasks related to hypertension (71%).

**Figure 2.**
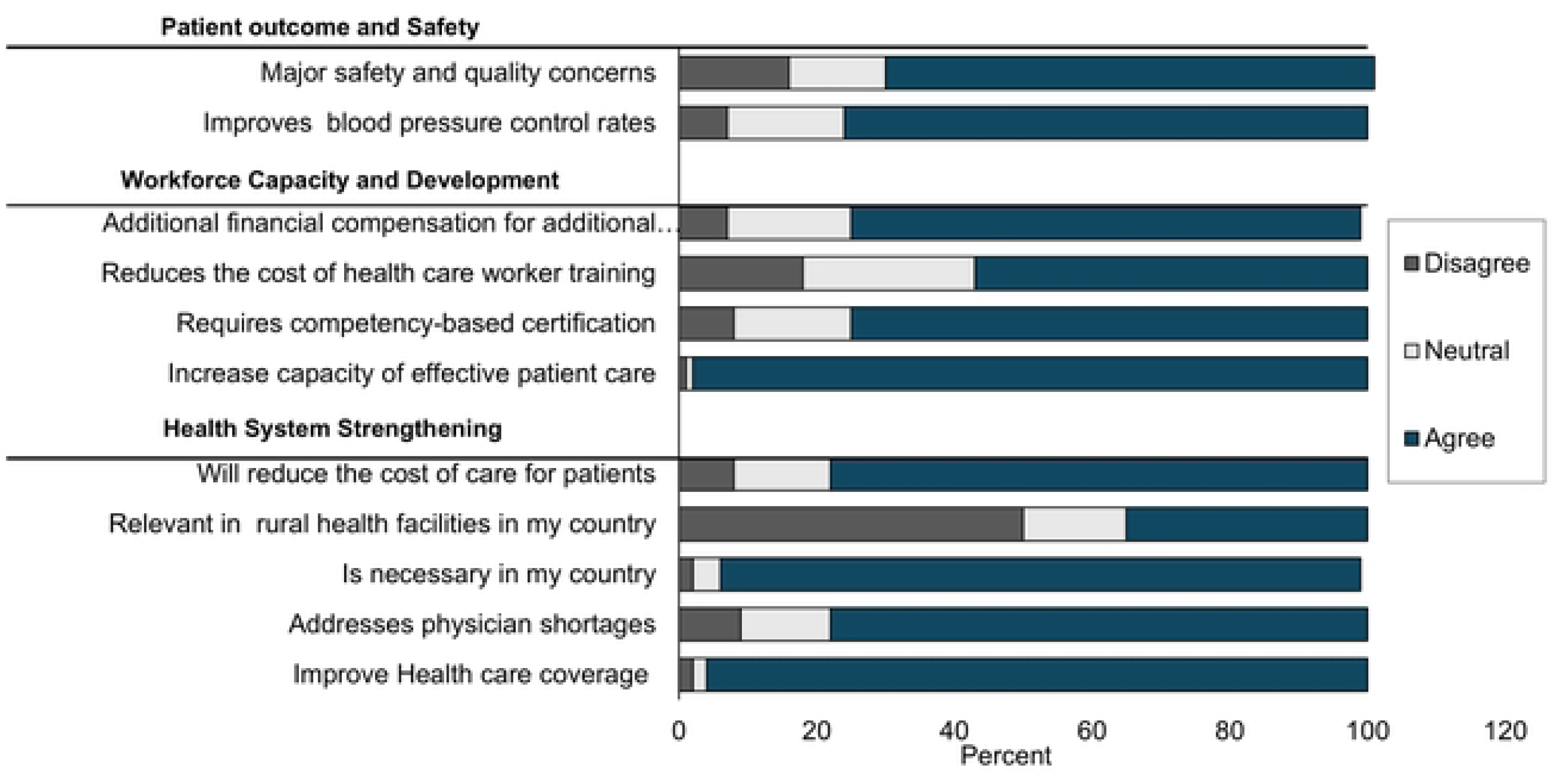
Perspectives on team-based care for hypertension management in Ghana

### Barriers and Facilitators for Team-Based Care

The major barriers to team-based care reported by HCWs were limited scope of practice laws and practice standards (76%) and inadequate training of non-physicians (74%). At least half of those indicated opposition by physicians (61%) and resistance by patients (51%) as challenges to the uptake of team-based care. Fewer than half (42%) suggested professional opposition from nurses (42%) (**Figure 3**). When barriers were stratified by HCW type, opposition by physicians was perceived as a more significant barrier compared to opposition by nurses. A comparatively higher proportion of HCWs reported physician opposition as a challenge: physicians (41%), nurses (63%), pharmacists (70%), physician assistants (88%), and community health nurses/workers (65%). In contrast, lower proportions reported opposition by nurses as a barrier: physicians (34%), nurses (44%), pharmacists (47%), physician assistants (46%), and CHN/CHWs (43%). There was relatively higher agreement across all cadres regarding the limited scope-of-practice laws and inadequate compensation for non-physician HCWs, compared to the level of agreement on patient resistance (**Online Supplemental Figure 4**).

**Figure 3.**
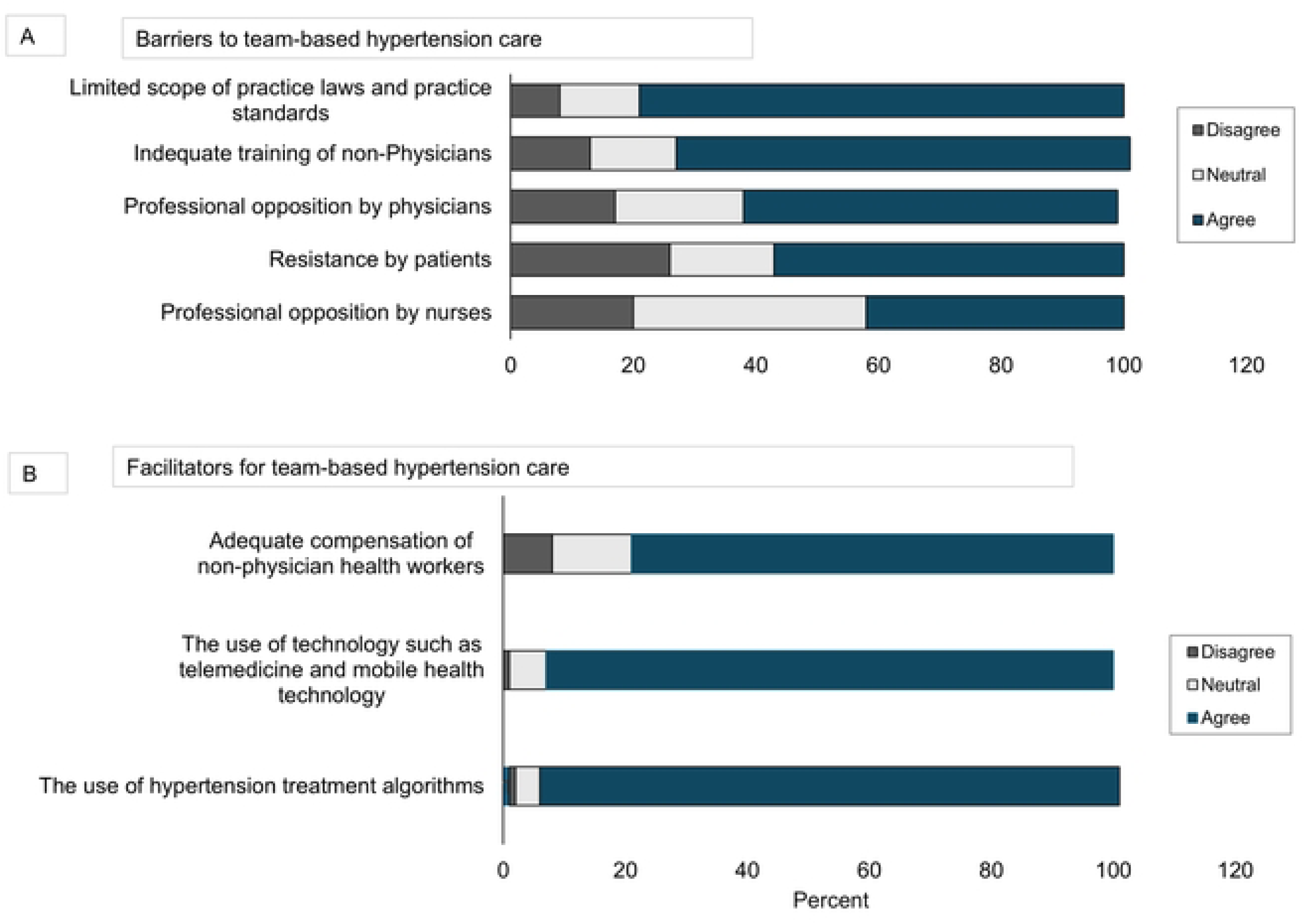
Barriers to team-based care in Ghana

Conversely, facilitators were identified, including the use of hypertension treatment algorithms (95%) and the use of technology (93%) by HCWs. Further, 79% of respondents indicated that adequate compensation for non-physician HCWs is a key enabler of team-based care in hypertension management (**Figure 4**). When responses were compared across HCW cadres, there was an even distribution of responses regarding what is considered a facilitator, except for the use of a hypertension treatment algorithm, where no physician, physician assistant, or CHN/CHW disagreed, except 3% of nurses (**Online Supplemental Figure 4**).

**Figure 4.** Facilitators for team-based care in Ghana

## DISCUSSION

This study is the first to assess team-based care practices for hypertension management across all 16 administrative regions in Ghana. Our findings provide an overview of task distribution among HCWs within the team-based care model, which is shaped by the broader task-shifting policy for non-communicable disease control. Guided by the framework of team-based care for hypertension management, we identified clear patterns of task-sharing across cadres, revealed critical barriers and facilitators to team-based care, and highlighted disparities in clinical task performance based on setting, experience, and education level.

Administrative tasks were primarily handled by non-clinical staff, such as records officers and data personnel, except for appointment scheduling, which was managed by physician assistants and physicians. Although this finding aligns with data from other LMICs where similar studies have been conducted[18], it contrasts with practices in high-income countries, where non-clinical team members typically manage scheduling. A potential explanation may be due to the arrangement of service delivery, which allows only prescribers (*mostly physicians and physician assistants*) to schedule appointments directly with their patients during consultations. Additionally, nurses and other non-physician HCWs were notably responsible for basic clinical duties.

Unsurprisingly, patient counseling was shared among various healthcare workers, though pharmacists were more frequently involved, likely due to the mandatory counseling required during medication dispensing. While some level of task-sharing characterized both administrative and basic clinical tasks, HCWs were seen mostly dominating their job specifications. However, establishing diagnosis and initiating treatment were mainly performed by physicians and physician assistants. This finding raises critical concerns for team-based care, given that long waiting times, driven by high patient volumes and physician shortages, stem from gaps in advanced clinical tasks.

Improving hypertension control at the facility level requires moving away from the traditional model of care to a team-based approach that allows HCWs to go beyond basic or administrative roles and take on advanced clinical responsibilities, which can significantly reduce delays, enhance patient-provider engagement, and support more holistic and coordinated care. Perhaps, reorganizing the hypertension care workflow to allow non-clinicians to schedule appointments may improve efficiency. This change would also go a long way toward serving as a gate-keeping system, where the existing workforce can effectively manage attendance at clinical visits. Additionally, when tasks were stratified by facility and HCW characteristics, hypertension tasks were more likely to be performed by HCWs with more years of experience, based in a district hospital (the highest level of primary healthcare), and in urban settings. However, those with lower levels of education mainly were seen performing administrative and basic clinical tasks rather than advanced clinical tasks. These findings highlight the need to tailor team-based care models by leveraging the expertise of more experienced HCWs in advanced clinical roles, while also developing targeted training programs to enable less-educated staff to take on expanded responsibilities.

Overall, HCWs across Ghana overwhelmingly perceive TBC as an essential strategy to augment patient care capacity, mitigate physician shortages, and improve both healthcare coverage and cost-efficiency. While this perception provides grounds for structured and practical integration of TBC into the mainstream hypertension care in Ghana, this outlook may be hindered by a few barriers identified by HCWs. Prominently identified challenges include restrictive scope-of-practice laws and inadequate training for non-physician providers, which collectively constrain non-physician HCWs from taking on advanced clinical tasks. Beyond barriers, professional opposition from physicians, alongside some resistance from patients, further complicates the implementation of team-based care. While the underpinning reason is beyond the scope of the current assessment, the majority of HCWs expressed concerns about patient safety and quality of care, perhaps when care is delivered by other HCWs, which may, in part, explain the observed opposition. Nonetheless, earlier studies from other LMICs have shown general acceptance across different cadres.

On the other hand, HCWs suggested practical recommendations to scale the implementation of team-based care for hypertension care in Ghana. Most of them agreed to the use of standardized hypertension treatment algorithms as a practical tool for guiding consistent care across diverse HCW teams. Moreover, using treatment algorithms in clinical settings has been shown to reduce variability in clinical decision-making, enhance confidence among non-physician providers, and promote adherence to evidence-based practices [19]. Beyond patient-centered benefits, use of technology in hypertension care should be seen as a strategic means to strengthen coordination, streamline communication, and foster more effective collaboration across care teams [10, 20].

Finally, adequate compensation for non-physician HCWs was identified, highlighting the need to incentivize expanded roles and recognize the increased responsibilities that come with team-based care[19]. Financial compensation for HCWs is a crucial motivational factor in healthcare workforce engagement in Ghana, following the implementation of the conditions of service policy, which allows facility-level remuneration of HCWs for all additional duties outside their job description. Indeed, for the fair and sustainable implementation of team-based care, there should be demonstrable commitment by both the Ministry of Health and facility leaders to compensate HCWs who perform additional tasks [21, 22]. This factor is critical for fostering the sustainability of facility-level implementation of team-based care and nurturing the commitment and motivation of HCWs.

### Limitations and Strengths

However, this study has some limitations. The use of a non-random sampling approach, which was essential for achieving broad geographical coverage, can introduce selection bias. Similarly, the use of online surveys may inherently exclude perspectives from individuals with limited digital or internet access, potentially leading to a response bias.

Nevertheless, this study has numerous strengths. (1) This study provides a unique and timely national perspective on TBC in Ghana, beyond localized observations, to offer a comprehensive understanding of current perspectives of team-based care for hypertension management in Ghana. (2) Most of the responses were from HCWs working at the highest level of primary healthcare, where most patients with hypertension receive care. (3) The responses received included at least a fair representation of representatives, and HCWs were predominantly involved in hypertension care at all levels of care in Ghana. (4) As the burden of hypertension continues to grow, there is an urgent need for policy implementers and healthcare leaders to move beyond rhetoric and policy development toward a stronger commitment to implementation of team-based care at the facility level. This shift is essential to overcoming the persistent bottlenecks that hinder hypertension control efforts in Ghana.

In conclusion, our findings have significant implications for hypertension management in Ghana. The findings strongly support the need for deliberate regulatory reforms to expand the legal scope of practice for non-physician HCWs. At the same time, strategic investments in targeted, competency-based training are essential to prepare these HCWs for more advanced clinical roles. For a more sustained gain, these efforts should be embedded within the broader structured task-shifting policy for the non-communicable disease control program of the Ministry of Health. More importantly, the findings resonate with the core framework of the WHO-HEARTS program, which offers a comprehensive model for implementing team-based care in LMICs[16].

## Data Availability Statement

Data are available upon reasonable request. Data will be made available upon request from researchers. Data will be de-identified and will strictly adhere to participant confidentiality and consent, per Institutional Review Board guidelines for each participating institution.

## Data Availability

Data cannot be shared publicly because it represent the perspective HCW workers and personal details. Data are available from the Institutional Data Access / Ethics Committee (contact via) for researchers who meet the criteria for access to confidential data.

## Acknowledgments

We thank the country representatives and healthcare workers who participated in this study.

## ABBREVIATIONS

HCW: Healthcare Worker
TBC: Team-Based Care
LMIC: Low- and Middle-Income Country
WHO: World Health Organization
RTSL: Resolve to Save Lives
STROBE: Strengthening the Reporting of Observational Studies in Epidemiology
BP: Blood Pressure
CHN: Community Health Nurse
CHW: Community Health Worker
NCD: Non-Communicable Disease
REDCap: Research Electronic Data Capture
GHS: Ghana Health Service

## ONLINE SUPPLEMENTAL MATERIAL

Appendix 1: Survey

See separate file.

## Supplemental Figure Legends

**Online Supplemental Figure 1**. Distribution of responses from HCWs across 16 regions

**Online Supplemental Figure 2**. Advanced clinic tasks stratified by facility and HCW characteristics

**Online Supplemental Figure 3**. Duration of antihypertensive medication refill by level of Blood pressure Control

**Online Supplemental Figure 4**. Facilitators to team-based care. (A) Agreement levels by topic; (B) Agreement levels by HCW cadre

**Online Supplemental Table 1.** Respondent Characteristics by Region

**Supplemental Table 1.** Respondent Characteristics by Region

| Online Supplemental Table 1. Respondent Characteristics by Region |  |  |  |  |  |  |  |  |  |  |  |  |  |  |  |  |
| --- | --- | --- | --- | --- | --- | --- | --- | --- | --- | --- | --- | --- | --- | --- | --- | --- |
|  | WNR | ASR | BOR | AHR | OTR | GAR | NOR | UWR | UER | SVR | VR | BER | CR | NER | ER | WR |
| Age, m (SD) | 31 (5) | 33 (4) | 33 (5) | 32 (6) | 34 (6) | 34 (9) | 36 (7) | 37 (4) | 34 (5) | 32 (4) | 32 (4) | 37 (7) | 37 (8) | 37 (2) | 36 (9) | 34 (7) |
| **Sex, n (%)** |  |  |  |  |  |  |  |  |  |  |  |  |  |  |  |  |
| Male | 12(70.6) | 16(50) | 13(43.3) | 10(100) | 11(47.8) | 16(38.1) | 34(69.4) | 8(88.9) | 4(66.7) | 10(76.9) | 9(42.9) | 6(60) | 10(66.7) | 4(100) | 4(57.1) | 6(66.7) |
| Female | 5(29.4) | 16(50) | 17(56.7) | 0 | 12(52.2) | 26(62.9) | 15(30.6) | 1(11.1) | 2(33.3) | 3(23.1) | 12(57.1) | 40(40) | 5(33.3) | 0 | 3(42.9) | 3(33.3) |
| **Profession, n (%)** |  |  |  |  |  |  |  |  |  |  |  |  |  |  |  |  |
| Physicians | 3(33.3) | 7(22.6) | 5(19.2) | 1(11.1) | 5(20.9) | 16(38.1) | 11(24.4) | 3(42.9) | 2(40.0) | 3(25.0) | 7(35.0) | 1(9.1) | 3(21.4) | 0 | 1(14.3) | 3(33.3) |
| Nursing Professionals | 5(55.6) | 12(38.7) | 13(50.0) | 5(55.6) | 16(66.7) | 15(35.7) | 24(53.3) | 1(14.3) | 2(40.0) | 4(33.3) | 6(30.0) | 6(54.6) | 1(7.1) | 1(33.3) | 5(71.4) | 5 (55.6) |
| Pharmacists | 0 | 2(6.5) | 0 | 0 | 0 | 3(7.1) | 4(8.9) | 0 | 0 | 0 | 4(20.0) | 2(18.2) | 10(71.4) | 1 (33.3) | 1(14.3) | 0 |
| Community Health Worker | 0 | 2(6.5) | 1(3.9) | 2(22.2) | 2(8.3) | 2(4.7) | 1(2.2) | 0 | 1(20.0) | 4(33.3) | 3(15.0) | 0 | 0 | 0 | 0 | 0 |
| Physician Assistants | 0 | 2(6.5) | 1(3.9) | 0 | 0 | 1(2.4) | 3(6.7) | 2(28.6) | 0 | 1(8.3) | 0 | 1(9.1) | 0 | 1(33.3) | 0 | 1(11.1) |
| Other<sup>a</sup> | 0 | 6(19.4) | 6(23.1) | 1(11.1) | 1(4.2) | 1(2.4) | 2(4.4) | 1(14.3) | 0 | 0 | 0 | 1(9.1) | 0 | 0 | 0 |  |
| **Education/Training, n (%)** |  |  |  |  |  |  |  |  |  |  |  |  |  |  |  |  |
| Diploma or lower | 6(37.5) | 6(20.7) | 11(35.5) | 6(66.7) | 8(36.4) | 9(23.1) | 9(20.0) | 0 | 0 | 3(30.0) | 7(36.8) | 4(40.0) | 0 | 1(25.0) | 2(28.6) | 5(55.6) |
| Bachelor's | 7(43.8) | 18(62.1) | 14(45.2) | 2(22.2) | 10(45.5) | 17(43.6) | 21(46.7) | 6(75.0) | 3(100) | 5(50.0) | 7(36.8) | 4(40.0) | 8(53.3) | 3(75.0) | 4(57.1) | 3(33.3) |
| Master's | 2(12.5) | 5(17.2) | 5(16.1) | 1(11.1) | 3(13.6) | 11(28.2) | 13(28.9) | 2(25.0) | 0 | 2(20.0) | 2(10.5) | 2(20.0) | 5(33.3) | 0 | 0 | 0 |
| Doctorate | 1(6.3) | 0 | 1(3.2) | 0 | 1(4.6) | 2(5.1) | 2(4.44) | 0 | 0 | 0 | 3(15.8) | 0 | 2(13.3) | 0 | 1(14.3) | 1(11.1) |
| **Workplace Setting** |  |  |  |  |  |  |  |  |  |  |  |  |  |  |  |  |
| Rural | 13(81.2) | 4(13.3) | 3(10.7) | 3(30.0) | 15(62.5) | 1(2.6) | 3(6.4) | 4(50.0) | 1(33.3) | 6(50.0) | 1(4.8) | 3(30) | 0 | 1(33.3) | 2(28.6) | 2(25.0) |
| Semi-Urban | 2(12.5) | 12(40.0) | 14(50.0) | 5(50.0) | 7(29.2) | 13(33.3) | 2(4.3) | 3(37.5) | 1(33.3) | 4(33.3) | 7(33.3) | 5(50.0) | 3(21.4) | 0 | 4(57.1) | 1(12.5) |
| Urban | 1(6.2) | 14(46.7) | 11(39.3) | 2(20.0) | 2(8.3) | 25(64.1) | 42(89.4) | 1(12.5) | 1(33.3) | 2(16.7) | 13(61.9) | 2(20.0) | 11(78.6) | 2(66.7) | 1(14.3) | 5(62.5) |
| **Facility Type, n (%)** |  |  |  |  |  |  |  |  |  |  |  |  |  |  |  |  |
| Tertiary Hospital | 0 | 2(11.8) | 3(17.7) | 0 | 2(13.3) | 3(14.3) | 19(90.5) | 0 | 3(60.0) | 0 | 7(63.6) | 0 | 3(60.0) | 0 | 0 | 0 |
| District Hospital | 6(100) | 13(76.5) | 13(76.5) | 4(50.0) | 12(80.0) | 7(33.3) | 1(4.8) | 1(20.0) | 2(40.0) | 4(80) | 3(27.2) | 3(75.0) | 2(40.0) | 13(92.9) | 3(75.0) | 6(100) |
| Lower Level | 0 | 2(11.8) | 1(5.9) | 4(50.0) | 1(6.7) | 11(52.4) | 1(4.8) | 4(80.0) | 0 | 2(20) | 1(9.1) | 1(25.0) | 0 | 1(7.1) | 1(25.0) | 0 |
| **Facility Telehealth Use, n (%)** |  |  |  |  |  |  |  |  |  |  |  |  |  |  |  |  |
| Used in primary care | 4(25.0) | 7(23.3) | 8(29.6) | 1(10.0) | 7(29.2) | 9(22.5) | 9(20.0) | 0 | 1(20.0) | 3(25.0) | 7(36.8) | 2(20.0) | 2(16.7) | 1(25.0) | 3(42.9) | 3(33.3) |
| Existence of Telehealth guideline | 3(75.0) | 6(85.7) | 5(62.5) | 0 | 3(42.9) | 5(55.6) | 3(42.9) | 0 | 1(100) | 2 (66.7) | 7(100) | 1(50.0) | 0 | 0 | 1(33.3) | 1(50.0) |

<sup>a</sup>This includes other community health centers, community clinics, and community pharmacy workers.
<sup>b</sup>Multiple choice question, answer choices total N=345.
SD: Standard Deviation;
Regions: Central (CER). Eastern (EAR). Greater Accra (GAR). North East (NER). Northern (NOR). Oti (OTR). Savannah (SAR). Upper East (UER). Upper West (UWR). Volta (VOR). Western (WER). Western North (WNR)

## Notes

### Competing Interest Statement

The authors have declared no competing interest.

### Funding Statement

Yes

### Author Declarations

This study was reviewed and approved by the Ghana Health Service Ethics Review Committee (Protocol ID: GHS-ERC026/03/23).

